# Hypertension in patients hospitalized with COVID-19 in Wuhan, China: A single-center retrospective observational study

**DOI:** 10.1101/2020.04.06.20054825

**Authors:** Zhenhua Zeng, Tong Sha, Yuan Zhang, Feng Wu, Hongbin Hu, Haijun Li, Jiafa Han, Wenhong Song, Qiaobing Huang, Zhongqing Chen

## Abstract

**Objectives:** It is unclear whether patients with hypertension are more likely to be infected with SARS-COV-2 than the general population and whether there is a difference in the severity of COVID-19 pneumonia in patients who have taken ACEI/ARB drugs to lower blood pressure compared to those who have not.

**Methods:** This observational study included data from all patients with clinically confirmed COVID-19 who were admitted to the Hankou Hospital, Wuhan, China between January 5 and March 8, 2020. Data were extracted from clinical and laboratory records. Follow-up was cutoff on March 8, 2020.

**Results:** A total of 274 patients, 75 with hypertension and 199 without hypertension, were included in the analysis. Patients with hypertension were older and were more likely to have pre-existing comorbidities, including chronic renal insufficiency, cardiovascular disease, diabetes mellitus, and cerebrovascular disease than patients without hypertension. Moreover, patients with hypertension tended to have higher positive COVID-19 PCR detection rates. Patients with hypertension who had previously taken ACEI/ARB drugs for antihypertensive treatment have an increased tendency to develop severe pneumonia after infection with SARS-COV-2 (P = 0.064).

**Conclusions:** COVID-19 patients with hypertension were significantly older and were more likely to have underlying comorbidities, including chronic renal insufficiency, cardiovascular disease, diabetes mellitus, and cerebrovascular disease. Patients with hypertension who had taken ACEI/ARB drugs for antihypertensive treatment have an increased tendency to develop severe pneumonia after infection with SARS-COV-2. In future studies, a larger sample size and multi-center clinical data will be needed to support our conclusions.

## Introduction

In December 2019, a cluster of severe cases of pneumonia of an unknown cause emerged in Wuhan, China, with clinical presentations greatly resembling viral pneumonia^1^ Deep sequencing analysis from lower respiratory tract samples indicated a novel coronavirus, which was named coronavirus disease (COVID-19). By Mar 16, 2020, the total number of patients had risen sharply to 153,546 confirmed cases worldwide (http://2019ncov.chinacdc.cn/2019-nCoV/global.html). The data extracted from 1099 patients with laboratory-confirmed COVID-19 in China on January 29, 2020 showed that hypertension is the most common coexisting illness in patients with COVID-19. Hypertension rates range from 13.4% in non-severe patients to 23.4% in severe patients, and the total incidence is as high as 15%^2^. Interestingly, angiotensin-converting enzyme 2 (ACE2) is involved in the pathogenesis of both hypertension and COVID-19 pneumonia. The above reasons prompted us to speculate that the organs attacked by SARS-COV-2 might not only be the lungs, but may also include other ACE2 expression organs, especially the cardiovascular system. In addition, patients with comorbid hypertension, especially those with long-term oral ACEI/ARB medication for hypertension, might have different susceptibility and severity levels of pneumonia upon infection with SARS-COV-2. Therefore, we investigated and compared the demographic characteristics, coexisting diseases, severity of pneumonia, and the effect of antihypertensive drugs (ACEI/ARB versus non-ACEI/ARB) in COVID-19 patients with coexisting hypertension. Our findings may hopefully reduce the mortality and morbidity associated with hypertension in patients with COVID-19.

## Methods

### Study design and participants

This single-center, retrospective, observational study was conducted at Hankou Hospital, which is one of the hospitals designated to treat COVID-19 pneumonia patients in Wuhan, China. The criteria for suspected and confirmed cases of COVID-19 were based on the criteria previously established by the WHO^3, 4^ and patients who met the WHO criteria were included in our study. Patients who were < 18 years, whose entire stay in hospital lasted for < 48 hours were excluded from the analysis. This study was approved by the National Health Commission of China and the Ethics Commission of Hankou Hospital (hkyy2020-005). The Ethics Commission of Hankou Hospital waived the requirement for obtaining informed consent.

We obtained epidemiological, demographic, clinical, laboratory, management, and outcome data from patients’ medical records using standardized data collection forms. Clinical outcomes were followed up to March 8, 2020. To analyze the relationship between stage of hypertension and clinical outcome, a minimum follow-up time of 14 days was required, including follow-up of patients who were discharged before March 8, 2020. If data were missing from the records or clarification was needed, the researchers also directly communicated with patients or their families to ascertain epidemiological and symptom data. All data were checked by two physicians (TS and HH). Initial investigations included a complete blood count, coagulation profile, and serum biochemical test (including renal and liver function, creatine kinase, lactate dehydrogenase (LDH), electrolytes, and 10 common respiratory pathogens). A description of the treatment procedures and measurements has been published previously^4^.

### Study Definitions

The definition and classification of hypertension were obtained from previously published literature^5^. The definitions of severe and non-severe pneumonia were obtained from published literature^6^.

### Chest computed tomography

All patients received a chest computed tomography (CT) scan on admission. All CT images were reviewed by two fellowship-trained cardiothoracic radiologists (GX and FW), with approximately 5 years of experience each, using the Electronic Medical Record System of Hankou Hospital. Images were reviewed independently, and final decisions were reached by consensus. In cases of disagreement between the two primary radiologist interpretations, a third fellowship-trained cardiothoracic radiologist with 10 years of experience (HL) adjudicated the final decision. No negative control cases were examined, and no blinding occurred. The radiologists evaluated the initial CT scans and assigned quantitative scores for the characteristics using a recently published scoring system^7^. The total CT score was the sum of lung involvement (5 lobes, score 1-5 for each lobe, range from 0 none to 25 maximum)^8^.

### Outcomes

The primary variable of interest was 28-day mortality (calculated from the day of admission). Secondary outcome variables included the severity of pneumonia, length of hospital stay, discharge rate from hospital, and hospitalization rate.

### Ethics approval

This study was approved by the National Health Commission of China and the Ethics Commission of Hankou Hospital (hkyy2020-005).

### Consent

The Ethics Commission of Hankou Hospital waived the requirement for obtaining informed consent.

### Statistical analysis

Statistical analysis was performed using SPSS software for Windows, version 20.0 IBM Corp., Armonk, NY, USA). Descriptive analyses were performed for demographic, clinical, and laboratory data. Bivariate analyses were used to assess the association of hypertension status and different parameters of interest. Continuous data, such as white blood cell count, were converted into categorical variables (normal, low, or high levels). Multivariate analysis was not performed due to the small sample size. The odds ratio (OR), obtained by logistic regression, was used to measure the strength of the association of each factor with hypertension. A *P* value *<0*.*05* was considered to indicate statistical significance. The date symptom onset was used as the starting date. We conducted a death certificate search of medical records to determine whether any patients who were alive on discharge died subsequently and included these data in the analysis when applicable.

## Results

During the study period, a total of 274 patients were admitted and met the case definition of clinically confirmed COVID-19 and the other inclusion criteria. The characteristics of the patients on admission, both overall and according to hypertension status are shown in **Table 1**. There were 75 patients with hypertension and 199 patients without hypertension. Compared to patients without hypertension, patients with hypertension were significantly older and more likely to have underlying comorbidities, including chronic renal insufficiency, cardiovascular disease, diabetes mellitus, and cerebrovascular disease. Patients with hypertension tended to have a longer time from onset to admission and have higher positive COVID-19 PCR detection rates.

**Table 1.**
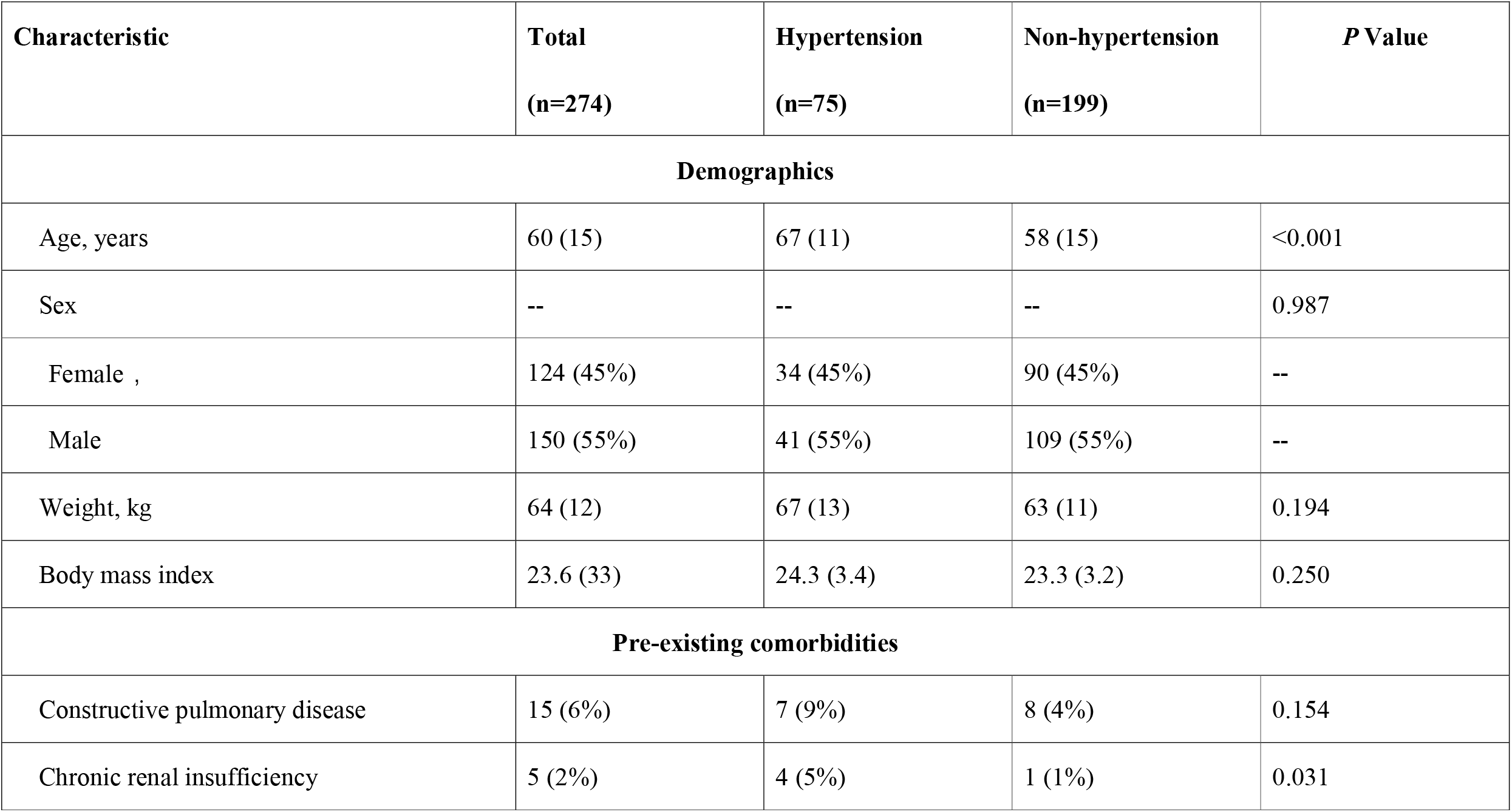
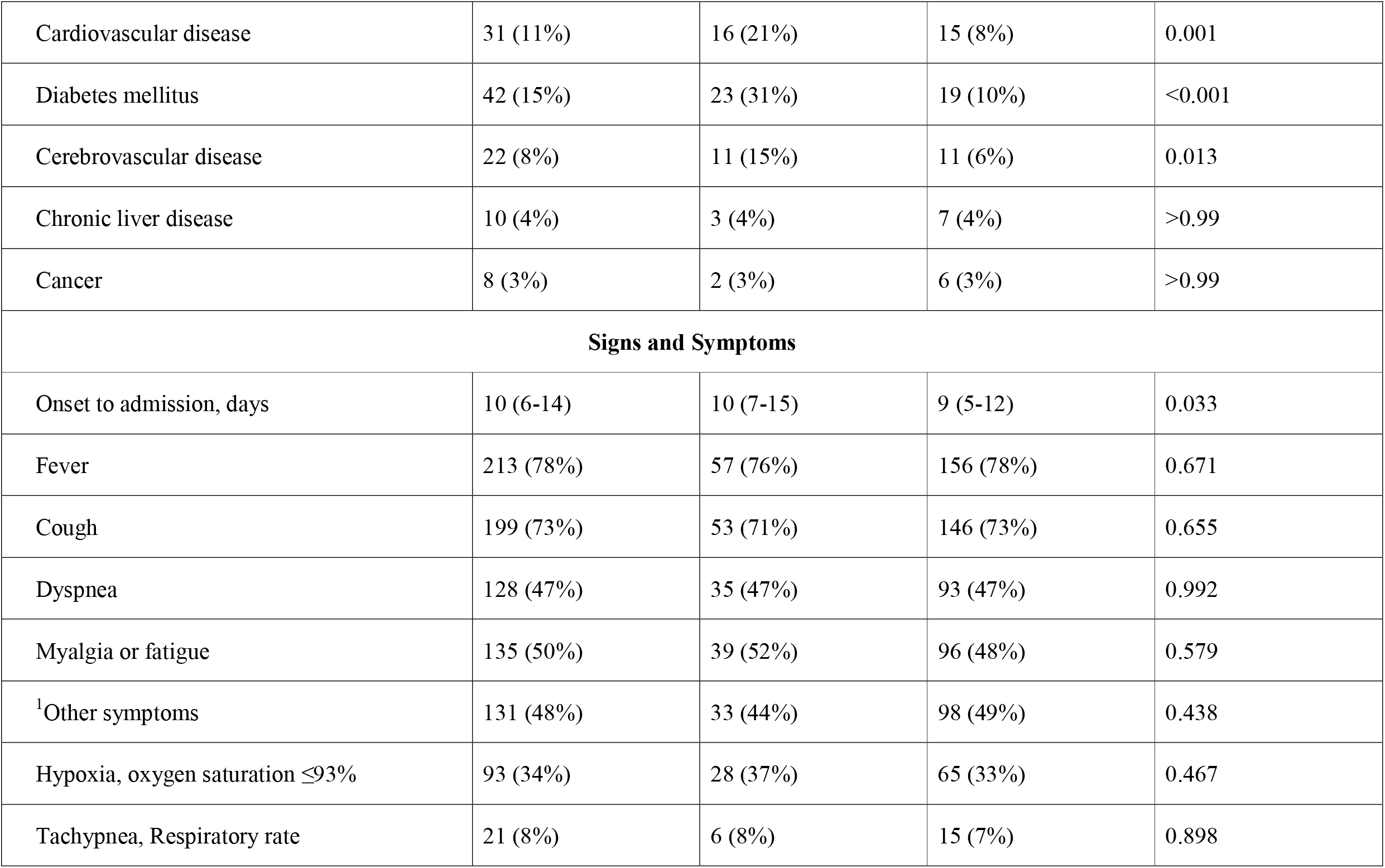
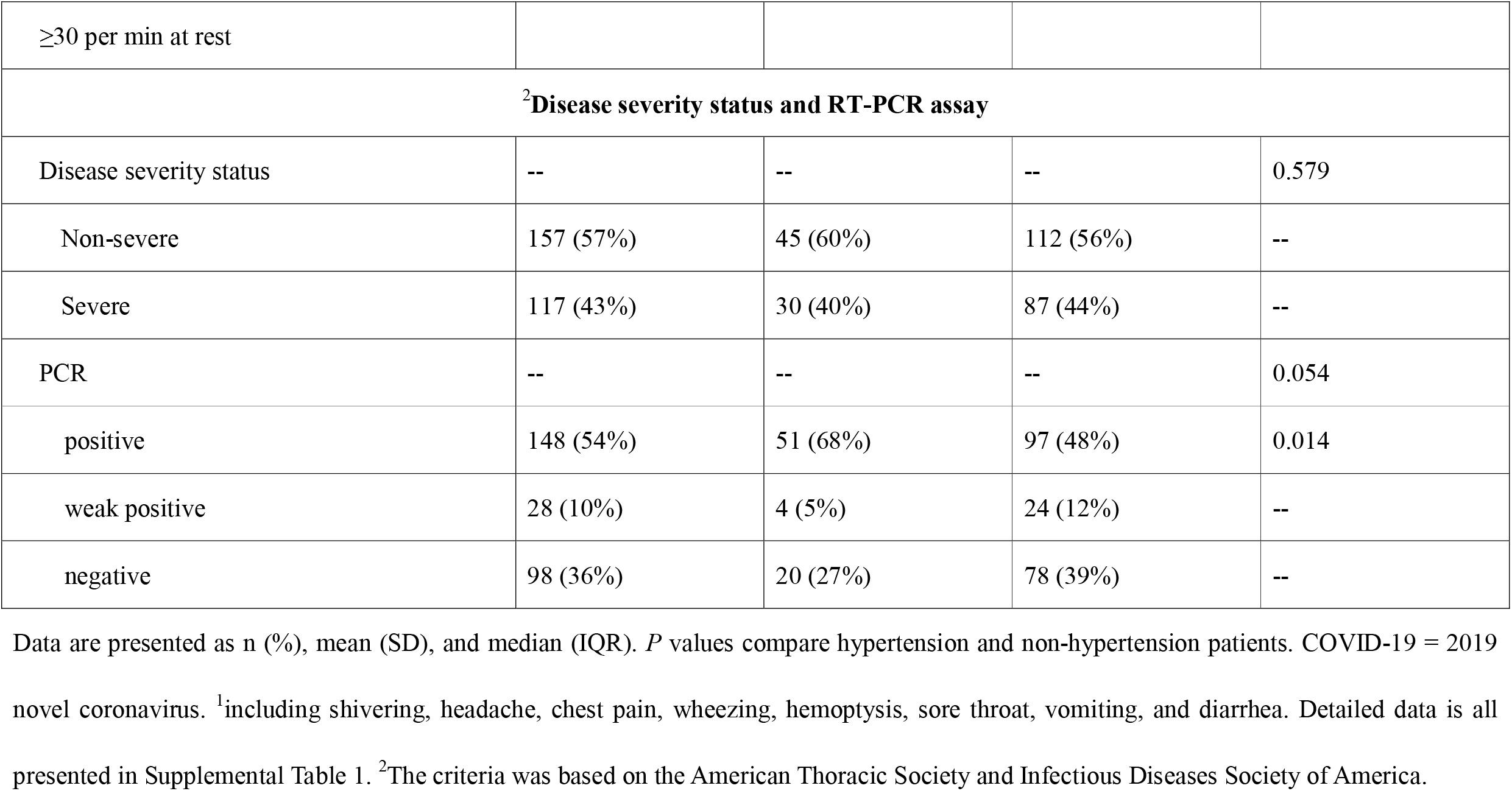
Demographics and clinical characteristics of patients with COVID-19 on admission.

**Table 2** shows the results of the laboratory tests performed on admission according to hypertension status. Patients with hypertension had higher levels of serum creatinine (SCr) and lower levels of hemoglobin and D-dimer in whole blood than patients without hypertension. No differences in clinical outcomes were found between patients with hypertension and without hypertension (**Table 3**).

**Table 2.** Laboratory findings of patients infected with COVID-19 on hospital admission.

|  | **Normal range** | **Total**<br>**(n = 274)** | **Hypertension**<br>**(n = 75 )** | **Non-Hypertension**<br>**(n = 199 )** | **P Value** |
| --- | --- | --- | --- | --- | --- |
| White blood cell count, $\times 10^9/\text{L}$ | 4.0-10.0 | 4.9 (3.8-7.1) | 5.3 (3.9-7.1) | 4.9 (3.6-7.2) | 0.231 |
| Neutrophil count, $\times 10^9/\text{L}$ | 1.8-6.3 | 3.6 (2.3-5.4) | 4.0 (2.4-6.4) | 3.4 (2.3-5.2) | 0.290 |
| Lymphocyte count, $\times 10^9/\text{L}$ | 1.1-3.2 | 0.8 (0.6-1.2) | 0.9 (0.6-1.2) | 0.8 (0.6-1.2) | 0.476 |
| Hemoglobin, g/L | 120-150 | 128 (17) | 124 (16) | 130 (17) | 0.011 |
| Prothrombin time, s | 11.0-16.0 | 14.7<br>(13.7-15.8) | 14.6<br>(13.3-15.7) | 14.8<br>(13.8-15.9) | 0.233 |
| Activated partial thromboplastin time, s | 27.0-45.0 | 35.3<br>(32.1-37.8) | 34.2<br>(32.1-37.4) | 35.4<br>(32.0-37.9) | 0.495 |
| D-dimer, mg/mL | 0.0-0.5 | 0.6 (0.1-2.0) | 0.5<br>(0.1-1.4) | 1.0<br>(0.2-3.3) | 0.011 |
| $\leq 0.5$ | | 129 (47%) | 26 (35%) | 103 (52%) | 0.024 |
| >0.5 to ≤1 |  | 40 (15%) | 11 (15%) | 29 (15%) | -- |
| >1 |  | 105 (38%) | 38 (50%) | 67 (33%) | -- |
| Alanine aminotransferase, U/L | 0-40 | 25 (17-37) | 24 (17-38) | 25 (16-37) | 0.878 |
| Aspartate aminotransferase, U/L | 0-40 | 27 (20-40) | 28 (21-44) | 27 (19-40) | 0.521 |
| Total bilirubin, μmol/L | 3.4-20.5 | 9.0 (6.6-12.0) | 8.6 (6.6-13.0) | 9.2 (6.4-11.8) | 0.959 |
| Albumin, g/L | 34.0-54.0 | 33.4 (4.9) | 32.6 (4.4) | 33.7 (5.0) | 0.096 |
| Creatinine, μmol/L | 50-120 | 69 (59-83) | 74 (61-95) | 68 (58-81) | 0.036 |
| Creatinine, μmol/L >120 |  | 16 (6%) | 9 (12%) | 7 (3%) | 0.017 |
| Creatine kinase, U/L | 30-310 | 100.<br>(58-167) | 103<br>(58-195) | 98<br>(58-160) | 0.594 |
| Lactate dehydrogenase, U/L | 120-150 | 251<br>(186-348) | 253<br>(185-344) | 251<br>(186-359) | 0.892 |
| Glucose (>11.1mmol/L) | 3.9-6.1 | 41 (15%) | 13 (17%) | 28 (14%) | 0.500 |
| Procalcitonin, ng/mL | <0.05 | 0.07 | 0.08 | 0.07 | 0.207 |

|  |  | (0.04-0.16) | (0.05-0.17) | (0.04-0.15) |  |
| --- | --- | --- | --- | --- | --- |
| <0.1 |  | 170 (62%) | 41 (55%) | 129 (65%) | 0.167 |
| ≥0.1 to <0.25 |  | 54 (20%) | 20 (26%) | 34 (17%) | -- |
| ≥0.25 to <0.5 |  | 25 (9%) | 5 (7%) | 20 (10%) | -- |
| ≥0.5 |  | 25 (9%) | 9 (12%) | 16 (8%) | -- |
| C-reactive protein, mg/L | ≤5.0 | 22.5<br>(4.0-37.3) | 21.7<br>(4.0-43.7) | 23.2<br>(4.0-37.0) | 0.845 |
| Hyperkalemia (>5.0mmol/L) | ≤5.0 | 21 (8%) | 7 (9%) | 14 (7%) | 0.531 |
| <sup>1</sup>Ten respiratory pathogens | NA | 3 (1%) | 1 (1%) | 2 (1%) | >0.99 |
| <sup>2</sup>CT score | NA | 7 (5-12) | 7 (5-13) | 7 (4-11) | 0.276 |

Data are presented as n (%), mean (SD), and median (IQR). *P* values compare hypertension and non-hypertension patients. COVID-19 = 2019 novel coronavirus.
<sup>1</sup>For each patient, the initial CT scans were evaluated and quantitatively scored for the characteristics referenced in recently published literature<sup>8</sup>.
<sup>2</sup>Including: influenza A virus, influenza B virus, rhinovirus/enterovirus, respiratory syncytial virus (RSV) A/B, adenovirus, mycoplasma
pneumoniae, parainfluenza viruses, *Bordetella pertussis*, *legionella pneumophila*, and *Chlamydophila pneumoniae*.

**Table 3.** Clinical outcomes of patients with COVID-19 at the data cutoff.

|  | **All patients**<br>**(n=274)** | **Hypertension**<br>**(n=75)** | **Non-Hypertension**<br>**(n =199)** | ***P* Value** |
| --- | --- | --- | --- | --- |
| Discharged from hospital | 215 (78%) | 63 (84%) | 152 (76%) | 0.098 |
| Hospitalization | 38 (14%) | 5 (7%) | 33 (17%) |  |
| Death | 21 (8%) | 7 (9%) | 14 (7%) |  |
| Length of hospital stay, days | 20 (14-27) | 22 (15-26) | 19 (13-28) | 0.381 |

Data are presented as n (%) and median (IQR).
*P* values compare hypertension and non-hypertension patients. *P* <0.05 was considered statistically significant.

We then compared the characteristics and clinical outcomes of patients with COVID-19 who had a history of hypertension and those who had previously taken ACEI/ARB drugs and those who had not. More cases of severe pneumonia were seen among patients who had taken ACEI/ARB drugs than the patients who had not, even though those who had taken ACEI/ARB drugs were younger than those who had not (Age, *P* = 0.055; Pneumonia severity status on admission, *P* = 0.064). No significant differences in other indexes were found between the two groups (**Table 4**).

**Table 4.**
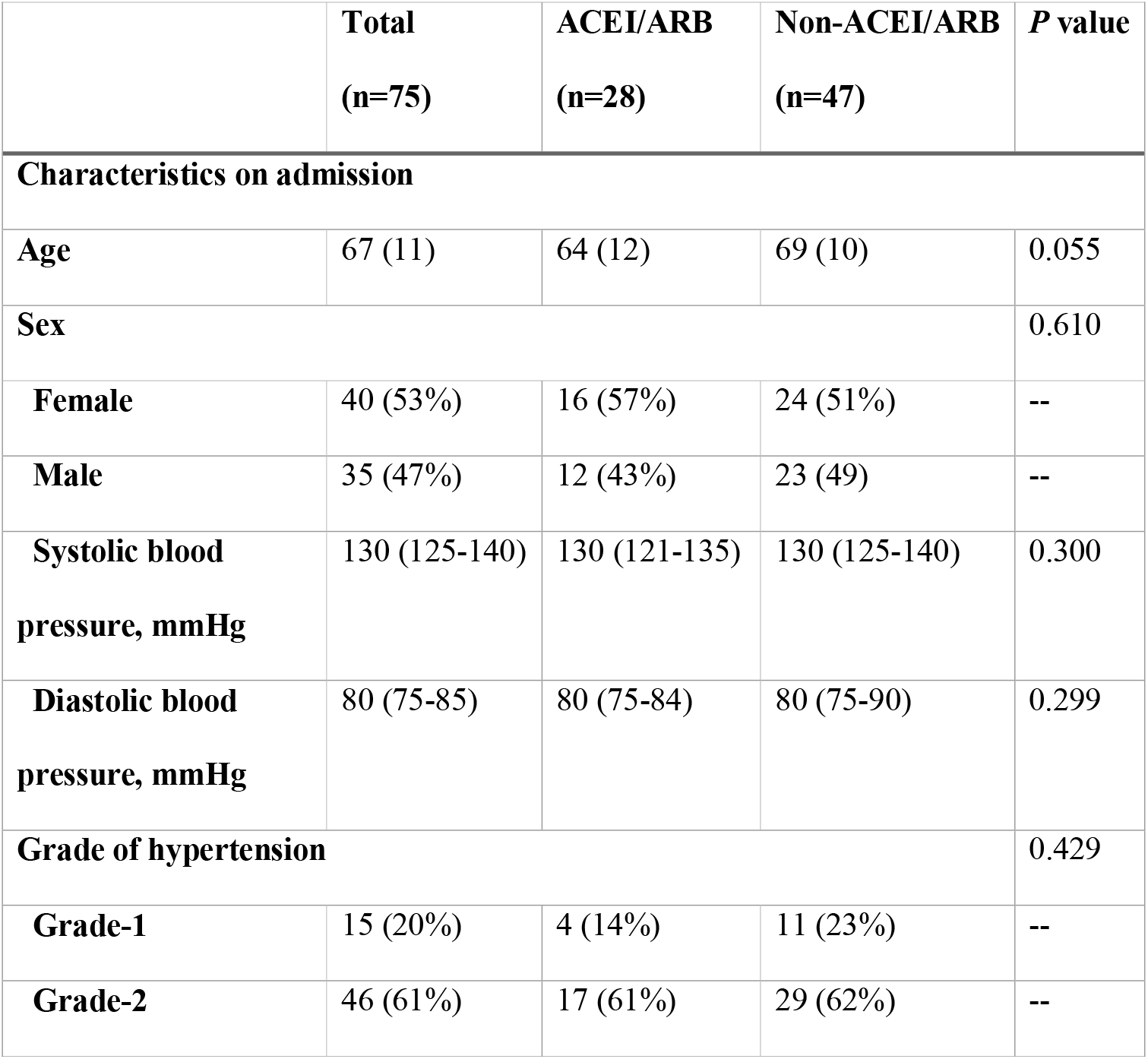
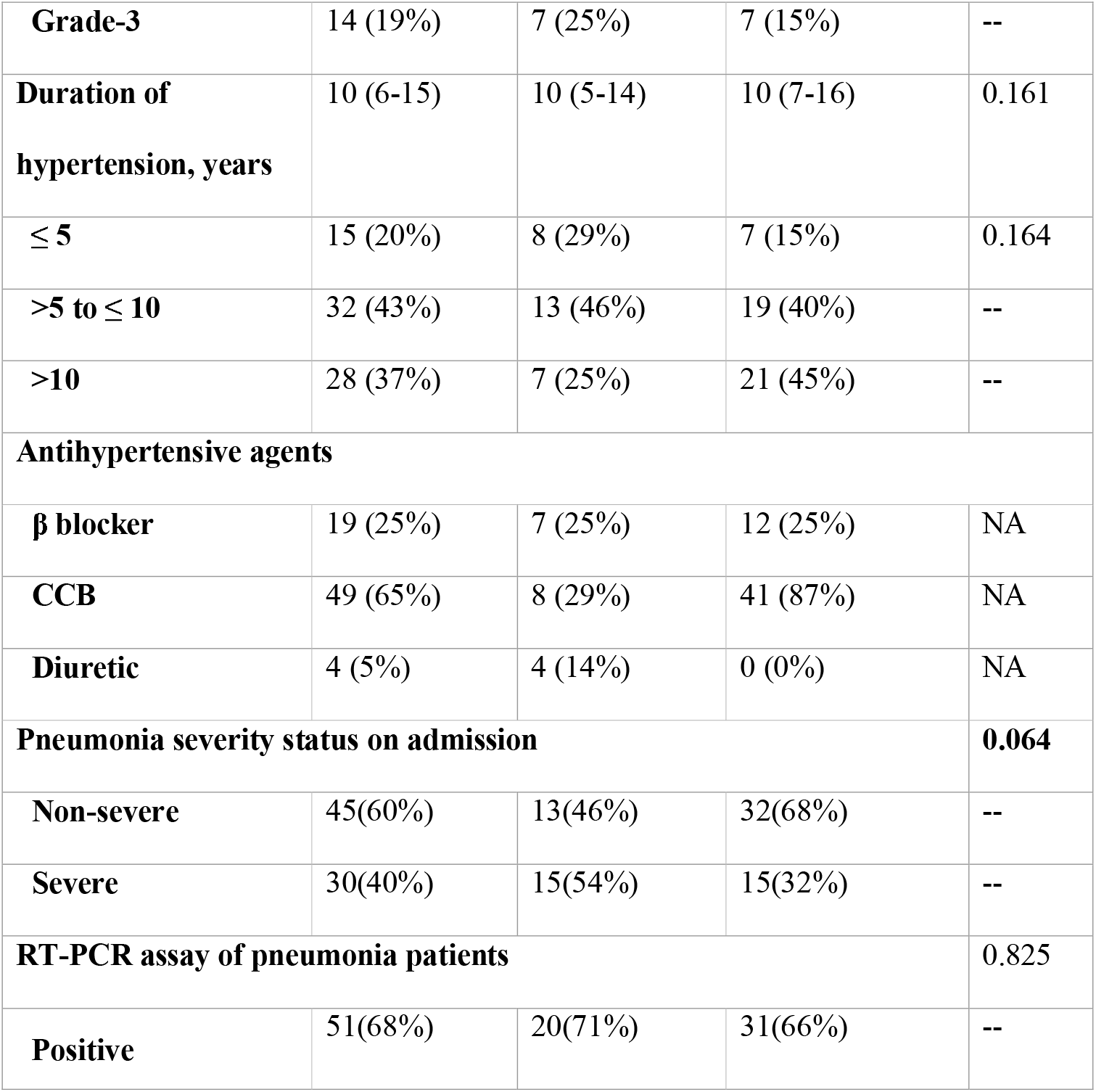
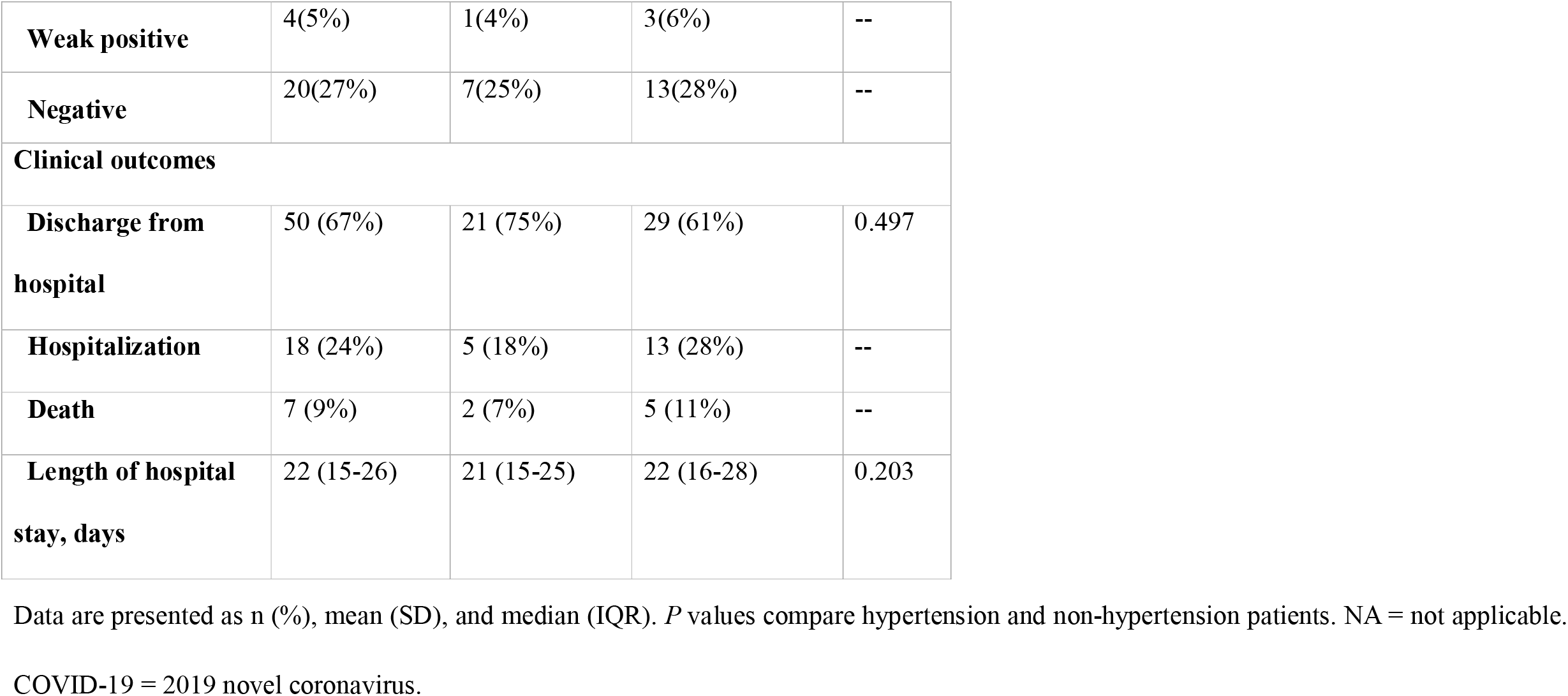
The characteristics and clinical outcomes of hypertension patients with COVID-19.

|  | **Total**<br>**(n=75)** | **ACEI/ARB**<br>**(n=28)** | **Non-ACEI/ARB**<br>**(n=47)** | **P value** |
| --- | --- | --- | --- | --- |
| **Characteristics on admission** |  |  |  |  |
| **Age** | 67 (11) | 64 (12) | 69 (10) | 0.055 |
| **Sex** |  |  |  | 0.610 |
| **Female** | 40 (53%) | 16 (57%) | 24 (51%) | -- |
| **Male** | 35 (47%) | 12 (43%) | 23 (49) | -- |
| **Systolic blood pressure, mmHg** | 130 (125-140) | 130 (121-135) | 130 (125-140) | 0.300 |
| **Diastolic blood pressure, mmHg** | 80 (75-85) | 80 (75-84) | 80 (75-90) | 0.299 |
| **Grade of hypertension** |  |  |  | 0.429 |
| **Grade-1** | 15 (20%) | 4 (14%) | 11 (23%) | -- |
| **Grade-2** | 46 (61%) | 17 (61%) | 29 (62%) | -- |
| **Grade-3** | 14 (19%) | 7 (25%) | 7 (15%) | -- |
| **Duration of hypertension, years** | 10 (6-15) | 10 (5-14) | 10 (7-16) | 0.161 |
| **≤ 5** | 15 (20%) | 8 (29%) | 7 (15%) | 0.164 |
| **>5 to ≤ 10** | 32 (43%) | 13 (46%) | 19 (40%) | -- |
| **>10** | 28 (37%) | 7 (25%) | 21 (45%) | -- |
| **Antihypertensive agents** |  |  |  |  |
| **β blocker** | 19 (25%) | 7 (25%) | 12 (25%) | NA |
| **CCB** | 49 (65%) | 8 (29%) | 41 (87%) | NA |
| **Diuretic** | 4 (5%) | 4 (14%) | 0 (0%) | NA |
| **Pneumonia severity status on admission** |  |  |  | **0.064** |
| **Non-severe** | 45(60%) | 13(46%) | 32(68%) | -- |
| **Severe** | 30(40%) | 15(54%) | 15(32%) | -- |
| **RT-PCR assay of pneumonia patients** |  |  |  | 0.825 |
| **Positive** | 51(68%) | 20(71%) | 31(66%) | -- |
| **Weak positive** | 4(5%) | 1(4%) | 3(6%) | -- |
| **Negative** | 20(27%) | 7(25%) | 13(28%) | -- |
| **Clinical outcomes** |  |  |  |  |
| **Discharge from hospital** | 50 (67%) | 21 (75%) | 29 (61%) | 0.497 |
| **Hospitalization** | 18 (24%) | 5 (18%) | 13 (28%) | -- |
| **Death** | 7 (9%) | 2 (7%) | 5 (11%) | -- |
| **Length of hospital stay, days** | 22 (15-26) | 21 (15-25) | 22 (16-28) | 0.203 |

Data are presented as n (%), mean (SD), and median (IQR). *P* values compare hypertension and non-hypertension patients. NA = not applicable.
COVID-19 = 2019 novel coronavirus.

## Discussion

Our observation shows that patients with hypertension who had previously taken ACEI/ARB drugs for antihypertensive treatment have an increased tendency to develop severe pneumonia after infection with SARS-COV-2. Hypertensive patients may need to adjust medications to be eliminate ACEI/ARB drugs to reduce the risk of severe pneumonia. Mechanistically, the increase in the zinc metallopeptidase ACE2 might help SARS-COV-2 enter the cells and accelerate its replication. ACE2 is the only known human homologue of the key regulator of blood pressure, angiotensin-converting enzyme (ACE). Since its discovery in 2000, ACE2 has been implicated to play an important role in the preservation of heart function, and the balance of blood pressure, with its effects being mediated, in part, through its ability to convert angiotensin II (Ang II) to angiotensin-(1–7). Unexpectedly, ACE2 also serves as the cellular entry point for the severe acute respiratory syndrome (SARS) virus and the enzyme is therefore a prime target for pharmacological intervention on coronavirus-related diseases^9^. In 2003, ACE2 was identified as the receptor for severe acute respiratory syndrome coronavirus (SARS-CoV), which mediates the infection and transmission of SARS-CoV. This effect does not rely on the protease activity on ACE^10^. Structural analysis showed that the spinous protein of SARS-CoV was in contact with the head-end of the ACE2 catalytic domain subunit I, did not involve subunit II, and did not block the active region. Once SARS-CoV is connected to ACE2, the extracellular part of ACE2 will be lysed, and the transmembrane part will be transferred into the cell, which mediates further fusion of the virus particles and host cells^11^. Through next-generation sequencing technology, the entire genome sequence of COVID-19 has been successfully determined^12^. Compared with the genomic sequences of previous coronaviruses, it is found that SARS-COV-2 has 79% similarity with SARS-CoV^13^.

It is worth noting that ACEI/ARB drugs that are administered to hypertensive patients have resulted in an increased expression of ACE2 in the lungs. The role of ACE2 is opposite to that of ACE. ACE2 can hydrolyze angiotensin I (Ang I) to produce angiotensin (1-7) peptides, thereby generating vasodilation and lowering blood pressure. ACEI/ARB drugs may increase the protein concentration of ACE2 in rat plasma, mRNA level of ACE2, and even the activity of ACE2 in cardiac tissue^14^. Thus, it is possible that those hypertensive patients who had been taking ACEI/ARB might have an increased level of mRNA and protein expression of ACE2 in the lungs, making them more vulnerable for SARS-COV-2 entry, as well as for development of severe COVID-19 pneumonia. These findings are worth investigating further.

However, some scholars believe that the down-regulation of ACE2 expression after a neo coronavirus challenge aggravates lung injuries. This opinion mainly comes from animal model studies in multiple laboratories and a small sample clinical study. Studies have shown that ACE2 expression decreases after viral infection, causing acute lung injury; exogenous supplementation with ACE2 could alleviate acute lung injury^15^. Human recombinant ACE 2 (rhuACE2) and angiotensin II type 2 receptors protect mice from severe acute lung injuries induced by acid aspiration or sepsis^16^. It was found that patients with acute respiratory distress syndrome (ARDS) who were treated with rhuACE2 injections had rapidly decreasing levels of Ang II and increasing levels of Ang1-7^17^. The down-regulation of ACE2 caused the imbalance of ACE/Ang II and ACE2 and the activity of Ang II in the renin-angiotensin-aldosterone system (RAAS) was enhanced because of the lack of antagonism^17^. Ang II mediates increased pulmonary vascular permeability by binding to receptor AT1, and might also cause pulmonary vascular contraction, resulting in hydrostatic pressure elevation and pulmonary edema^10, 17^. More clinical evidence is required to determine whether the expression of ACE2 is down-regulated in humans after a coronavirus (including COVID-19) infection.

The importance of this RAAS activation in the development of pulmonary edema in COVID-19 is still unknown. First, viral infections themselves, including COVID-19, may directly cause the destruction of alveolar type I and type II epithelial cells, and the alveolar-capillary membrane may be insufficient. Second, excessive inflammatory reactions due to the infection may also directly damage the integrity of the pulmonary capillary endothelial barrier, resulting in a massive increase of pulmonary vascular permeability and pulmonary edema, which appears as ground glass shadow on a chest CT^18^. If so, is it unknown whether ACE2 downregulation plays a key role in the impact of pulmonary edema and disease severity.

Nevertheless, the results of our study suggest that in the widespread COVID-19 pandemic, the hypertensive population may need to quickly consider their choice of antihypertensive medications to reduce the possibility of developing severe pneumonia. Our investigation found that COVID-19 patients taking ACEI/ARB agents are younger than patients taking non-ACEI/ARB agents; this may be related to their doctor’s prescribing habits. However, we cannot directly conclude that COVID-19 patients taking ACEI/ARB antihypertensive drugs will have more severe pneumonia than those not taking ACEI/ARB drugs because many confounding factors such as coexisting diseases have not yet been excluded.

Our survey shows that the population with hypertension does not seem to be more susceptible to COVID-19 comparing with the non-hypertension population. Previously, a large sample study of 1099 patients showed that hypertension is a coexisting disease in only approximately 15% cases of COVID-19^2^. Our study revealed that 27.4% (75/274) of the COVID-19 patients had hypertension, while the median age of patient sample we surveyed was generally older than that of previous reported populations^2^. Moreover, a previous nationwide survey in China from October 2012 to December 2015 showed that the overall rate of hypertension was 23.2% in Chinese adult population (≥18 years)^5^.

Our research has the following limitations: First, our survey only focused on Hankou Hospital in Wuhan, and the number of patients included was small; larger sample size and multi-center clinical data will be needed to support our conclusions in a future study. Second, our investigation included all clinically confirmed cases. Many patients were not tested because of the lack of nucleic acid test kits at the beginning of the outbreak. Further, there were some patients were suspected to have COVID-19 but were excluded from our study because they tested negative in the hospital; however, based on the exact epidemiological history, the patient’s signs and symptoms, and chest CT examination results, we trust the reliability of the COVID-19 diagnosis in these patients. Third, due to the urgent data collection, we had a shorter follow-up time for some patients. The shortest hospital stay for patients is only > 14 days. This may interfere with the final prognosis and lead to the failure to analyze survival times. Fourth, we did not analyze the comorbidities such as shock and ARDS, because our focus is to analyze the risk associated with hypertension to provide evidence for the issue of COVID-19 prevention and treatment guidelines.

### Perspectives

COVID-19 patients with coexisting hypertension were significantly older and were more likely to have underlying comorbidities, including chronic renal insufficiency, cardiovascular disease, diabetes mellitus, and cerebrovascular disease. Patients with hypertension tended to have a longer time from onset to admission and have higher positive COVID-19 PCR detection rates. Patients with hypertension who had taken ACEI/ARB drugs for antihypertensive treatment have an increased tendency to develop severe pneumonia after infection with SARS-COV-2. In future studies, a larger sample size and multi-center clinical data will be needed to support our conclusions.

## Data Availability

None

## Abbreviations and symbols

COVID-19: Coronavirus disease
ACE2: Angiotensin-converting enzyme 2
LDH: Lactate dehydrogenase
OR: Odds ratio
CT: Computed tomography
ARDS: Acute respiratory distress syndrome
RAAS: Renin-angiotensin-aldosterone system
SCr: Serum creatinine
Ang II: Angiotensin II
SARS: Severe acute respiratory syndrome
SARS-CoV: Severe acute respiratory syndrome coronavirus
Ang I: Angiotensin I
rhuACE2: Human recombinant ACE 2

## Acknowledgments

This work was supported by the Natural Science Foundation of China Grant 81871604; the Natural Science Foundation of Guangdong Province, China, Grants 2016A030310389 and 2017A030313590; the Outstanding Youths Development Scheme of Nanfang Hospital, Southern Medical University, Guangzhou, China, Grant 2016J011.

